# Impact of a SARS-COV-2 Infection in Patients with Celiac Disease

**DOI:** 10.1101/2020.12.15.20248039

**Authors:** Luca Elli, Federica Facciotti, Vincenza Lombardo, Alice Scricciolo, David S Sanders, Valentina Vaira, Donatella Barisani, Maurizio Vecchi, Andrea Costantino, Lucia Scaramella, Bernardo dell’Osso, Luisa Doneda, Leda Roncoroni

## Abstract

**Objective:** The SARS-CoV-2 pandemic has spread across the world causing a dramatic number of infections and deaths. No data are available about the effects of an infection in patients affected by celiac disease (CD) in terms of the development of related symptoms and antibodies. We aimed to investigate the impact of the SARS-CoV-2 pandemic in celiac patients.

**Design:** During a lockdown, the celiac patients living in the Milan area were contacted and interviewed about the development of COVID-19 symptoms as well as adherence to an anti-virus lifestyle and a gluten-free diet (GFD). They were also given a stress questionnaire to fill in. The development of anti-SARS-CoV-2 IgG and IgA (anti-RBD and N proteins) and the expression of the duodenal ACE2 receptor were investigated. When available, duodenal histology, anti-tissue transglutaminase IgA (tTGA), presence of immunologic comorbidities and adherence to the GFD were analysed as possible risk factors.

**Results:** 362 celiac patients have been interviewed and 42 (11%) presented with COVID-19 symptoms. The presence of symptoms was not influenced by tTGA positivity, presence of duodenal atrophy or adherence to GFD. 37% of the symptomatic patients presented anti-SARS-CoV-2 immunoglobulins (Ig). Globally, 18% of celiac patients showed anti-SARS-CoV-2 Ig vs 25% of the non-celiac control (p=0.18). The values of anti-RBD IgG/IgA and anti-N IgG did not differ from the non-celiac controls. Celiac patients had a significant lower level of anti-N IgA. The ACE2 receptor was detected in the non-atrophic duodenal mucosa of celiac patients; atrophy was associated with a lower expression of the ACE2 receptor.

**Conclusion:** CD patients have an anti-SARS-CoV-2 Ig positiveness and profile similar to non-celiac controls, except for anti-N IgA. The main celiac parameters and adherence to the GFD do not influence the development of a different Ig profile.

**What is already known about this subject?:** The SARS-CoV-2 pandemic has spread across the world causing infections and deaths. little is known about the possible relationship between autoimmune comorbidities and SARS-CoV-2 infection and COVID-19, and nothing it known about celiac disease.

**What are the new findings?:** In a large cohort of celiac patients living in a high SARS-CoV-2 incidence area in Northern Italy, no difference was observed evidenced in terms of the development of anti-SARS-CoV-2 Ig and their IgG and IgA profile compared with the normal population

**How might it impact clinical practice in the foreseeable future?:** The absence of a relationship between celiac disease and SARS-CoV-2/COVID-19 has a relevant impact on health policy to control the pandemic by supporting an optimal resource location.

## INTRODUCTION

Starting in January 2020, the dramatic SARS-CoV-2 (Severe Acute Respiratory Syndrome Coronavirus 2) pandemic has spread across the world reaching a dramatic number of infections and deaths (https://covid19.who.int/).[1]

The clinical picture of COVID-19 (Coronavirus disease 2019) is variable, ranging from an asymptomatic course to a severe form of pulmonary distress syndrome with high mortality rates. The most frequent symptoms reported in case of COVID-19 are fever, cough and dyspnoea but gastrointestinal symptoms have also been described.[2]

It is still unclear which factors influence the prognosis of COVID-19 patients, but it seems that comorbidities (e.g. hypertension or cardiologic disorders, overweight), smoking habits and male sex can be considered as risk factors; there is still doubt as to whether the presence of any immunological/autoimmune disorder can also be considered as a risk factor, but the preliminary data seem reassuring.[3–7]

According to the WHO guidelines, COVID-19 diagnosis is confirmed by the reverse transcription polymerase chain reaction (RT-PCR), performed on samples taken from the respiratory tract, e.g. nasopharyngeal swabs; it should be noted that SARS-CoV-2 RNA can be found in other specimens from COVID-19 patients, such as their stools.[8]

SARS-CoV-2 enters human cells using the ACE2 (Angiotensin Converting Enzyme 2) receptor, which is widely expressed on pneumocytes but also on other cells, such as enterocytes. This can explain the gastrointestinal tract involvement and, in particular, the small bowel (SB), where the relevant autoimmune diseases develop.[9,10]

Celiac Disease (CD) is one of the most common autoimmune disorders, affecting approximately 1% of the general population. It is triggered by the ingestion of gluten-containing food in genetically susceptible subjects carrying the HLA DQ2 and/or DQ8 haplotypes. CD is characterised by the serological presence of autoantibodies (tTGA – anti-tissue transglutaminase, IgA and IgG), duodenal villous atrophy, increased intra-epithelial lymphocytes (IELs) and crypt hyperplasia. The mainstay of treatment of CD is a gluten free diet (GFD), which generally leads to a good control of symptoms a complete recovery of duodenal stricture and a good prognosis. In a small percentage of cases, CD can encounter complications such as refractory celiac disease (RCD, subdivided into type I and II), ulcerative jejunoileitis, or Enteropathy-associated T-cell lymphoma (EATL).[11,12]

The possibility of any interaction between a SARS-CoV-2 infection and the immune system of CD subjects could be clinically and epidemiologically relevant. Nowadays, limited data and rather few indications exist regarding this important issue.[6,13] With the present study, we aimed to investigate the presence of a SARS-CoV-2 infection and COVID-19 among an Italian cohort of patients with CD living in an area (Milan) heavily hit by the pandemic.

## METHODS

### Patients

From March 8th 2020 to May 4th 2020, due to the COVID-19 lockdown, Italian outpatient clinics were closed for all non-urgent practice.[14] From 20 April 2020 to 30 June 2020, CD patients usually attending the “Center for the Prevention and Diagnosis of Celiac Disease”, Fondazione IRCCS Ca’ Granda Ospedale Maggiore Policlinico in Milan, were enrolled in a prospective monocentric study. Adult patients underwent a medically-assisted phone interview after they had given their oral informed consent. The following data were collected: date of birth, sex, presence of COVID-19 symptoms (flu-like symptoms, fever or chills, cough, shortness of breath or difficulty breathing, fatigue, muscle or body aches, headache, recent loss of taste or smell, sore throat, congestion or runny nose, nausea or vomiting and diarrhoea), the presence of a cohabitant with COVID-19, the execution of a SARS-CoV-2 PCR nasopharyngeal swab, adherence to an anti-virus lifestyle (shielding and social distancing, numerical rate scale from 0 to 10, NRS) and with GFD (NRS). When available within the last 12 months, duodenal histology findings (Marsh-Oberhuber classification),[15] anti-tissue transglutaminase IgA (tTGA) levels, presence of immunologic comorbidities and adherence to the GFD (nutritional assessment and urine gluten peptide detection) were analysed.

All the patients completed phone validated questionnaires: the ISMA (International Stress Management Association) Stress questionnaire [16]. The Stress questionnaire was used to evaluate the susceptibility of subjects to stress through a 25-item scale. It presents dichotomous response options: “yes” or “no” with a “yes” response corresponding to 1, while a “no” response corresponding to 0. A total score of 4 or lower means that there is a low probability that the patient is suffering of a stress-related illness; a score of 5-13 points is associated with more stress-related health, mental or physical problems, for which the patient could benefit from counselling; a score of 14 or more shows an inclination to stress with unhealthy behaviours.

This study was approved by the local Ethical Committee (reference number 458_2020).

### Patient and public involvement

Patients undergoing treatment at the “Centre for Prevention and Diagnosis of Celiac Disease” were consulted by way of phone calls and interviews and throughout the development of the study protocol. All the relevant feedback was considered and incorporated into patient information and consent sheets. During the ethical Committee consultation, a representative of the patients discussed the protocol and the consents. If possible, the results will be disseminated to all participants and to other health care professionals at scientific conferences and through press releases.

### Antigen proteins production

The recombinant Spike SARS-CoV-2 glycoprotein receptor binding domain (RBD) and the Nucleocapsid proteins have been produced in the COVID-19 laboratory of the European Institute of Oncology, IEO, by Drs Marina Mapelli and Sebastiano Pasqualato.

The RBD proteins have been produced in mammalian HEK293F cells as glycosylated proteins by transient transfection with pCAGGS vectors generated in Prof. Krammer’s laboratory [17]. The constructs were synthesised using the genomic sequence of the isolated virus, Wuhan-Hi-1 released in January 2020, and contain codons optimised for expression in mammalian cells. Secreted proteins were purified from the culture medium by affinity chromatography and quantified.

The recombinant nucleoprotein-N were produced in BL21-pLysS bacterial cells by pET28 overexpression vectors. The purification protocol comprised a first affinity chromatographic column, followed by a size exclusion column.

Retrieved proteins were quantified, flash frozen in liquid nitrogen in aliquots and stored at −80°C.

### Detection of anti-SARS-CoV-2 antibodies by ELISA

The ELISA assay to detect immunoglobulins (Ig) G and A uses a fragment of the SARS-CoV-2 Spike glycoprotein (S-protein) and the Nucleocapsid (N protein) as antigens based on the recently published protocol [17–19]. Briefly, after binding the proteins (RBD and N proteins) to a Nunc Maxisorp ELISA plate, and blocking aspecific bindings with PBS-BSA 3%, patients’ sera to be analysed were applied to the plate to allow antibody binding at a final dilution of 1:200, revealed with secondary anti-human-IgG (BD, clone G18-145) and IgA (Biolegend, Poly24110) antibody conjugated to HRP. Samples are read on a Glomax reader at 450 nm. This ELISA test is not intended for commercial use and it is currently under evaluation at the Italy’s Ministry of Health (Aut.Min.Rich. 15/05/2020) for emergency use approval.

Positivity threshold levels were determined by ROC curves. Positivity for RBD was OD 0.29 for IgG and 0.5 for IgA. Positivity for Nucleocapsid was OD 0.32 for IgG and 0.38 for IgA.

A sex aged match group of 164 non-CD subjects, tested in the same time frame of CD patients, has been used as the control.

### Histology and immunostaining

ACE2 immunostaining (bs-1004R; Bioss Antibodies) was performed on the endoscopic duodenal biopsies from CD patients using an automated immunostainer (Ventana Benchmark Ultra, Roche Diagnostics) as previously described [20]. ACE2 staining was scored as the percentage of positive enteric cells (0-100%) multiplied the intensity of the staining (0-3, where 0 stands for the absence of staining and 3 stands for intense staining). Therefore, the ACE2 score ranged from 0 to 300.

### Statistical analysis

The data are described as meanLJ±LJSD or median (interquartile range) unless otherwise indicated. The continuous demographic variables were compared between the groups using an independent Student’s *t*-test. Fisher’s exact test and Yate’s correction continued to be used to evaluate the distribution of categorical variables. A 5% Significance level was used, and the software packages STATA® v. 13.1 (StataCorp LLC, College Station, TX, USA) and GraphPad Prism v. 6 (GraphPad Software, La Jolla, CA, USA) were used for analysis and graph processing.

## RESULTS

### Patients and Symptoms

During the COVID-19 lockdown, 362 CD patients (288 (80%) females, age at enrolment 45±15 years, age at diagnosis 33±16) were called by phone in order to evaluate the occurrence of symptoms suggestive of COVID-19 or of a nasopharyngeal swab positive for SARS-CoV-2 infection. Among them, 21 (6%) had a diagnosis of refractory CD (RCD) according to international criteria. The following data were available: 202 (56%) patients underwent a urinary detection of gluten immunogenic peptides (uGIP), and in 23 of them (11%), the test detected gluten peptides in their urine; in 333 (91%) cases, tTGA values were available and indeed 78 (23%) patients showed a positive serology. In 201 (55%) patients, a recent duodenal histology was available. Globally, 42 (12%) patients reported symptoms potentially related to COVID-19 during the phone call. In figure 1, the percentage of referred symptoms is shown. In table 1, the clinical and demographic data of these CD patients are reported. Between CD patients with or without COVID-19 like symptoms, no statistically significant difference was found about age, age at CD diagnosis, sex, presence of tTGA or duodenal atrophy, GFD adherence or the presence of other autoimmune comorbidities. Only one patient had been hospitalised for mild respiratory distress without the need of intensive care or respiratory artificial support. Almost all patients reported a strict adherence to an antiviral lifestyle (NRS 9.7±0.6) and no one reported difficulties in the adherence to the GFD (NRS 9.7±0.9).

**Table 1.** Clinical and demographic characteristics of celiac patients with or without COVID-19 like symptoms

|  | Overall<br>N=362 | COVID-19 symptoms |  |  |
| --- | --- | --- | --- | --- |
|  |  | Present<br>N=42 | Absent<br>N=320 | P<br>(Present vs absent) |
| **Sex, female (%)** | 305(79) | 36(85) | 269(84) | 1.0 |
| **Age at enrolment (years)** | 45±15 | 41±13 | 46±15 | 0.05 |
| **Age at diagnosis (years)** | 32±15 | 30±13 | 34±16 | 0.13 |
| **BMI (kg/cm<sup>2</sup>)** | 22.4±3.4 | 22.2±3.6 | 22.3±3.9 | 0.96 |
| **Positive anti-transglutaminase IgA (%)** | 23 | 23 | 21 | 0.69 |
| **Nonadherence to GFD and/or positive urinary GIP (%)** | 10 | 10 | 11 | 1.0 |
| **Duodenal atrophy (Marsh 3a, 3b, 3c) (%)** | 64 | 59 | 67 | 0.47 |
| **Refractory celiac disease (%)** | 5 | 0 | 6 | 0.15 |
| **Presence of an AI comorbidity (%)** | 19 | 19 | 22 | 0.69 |

**Figure 1.**
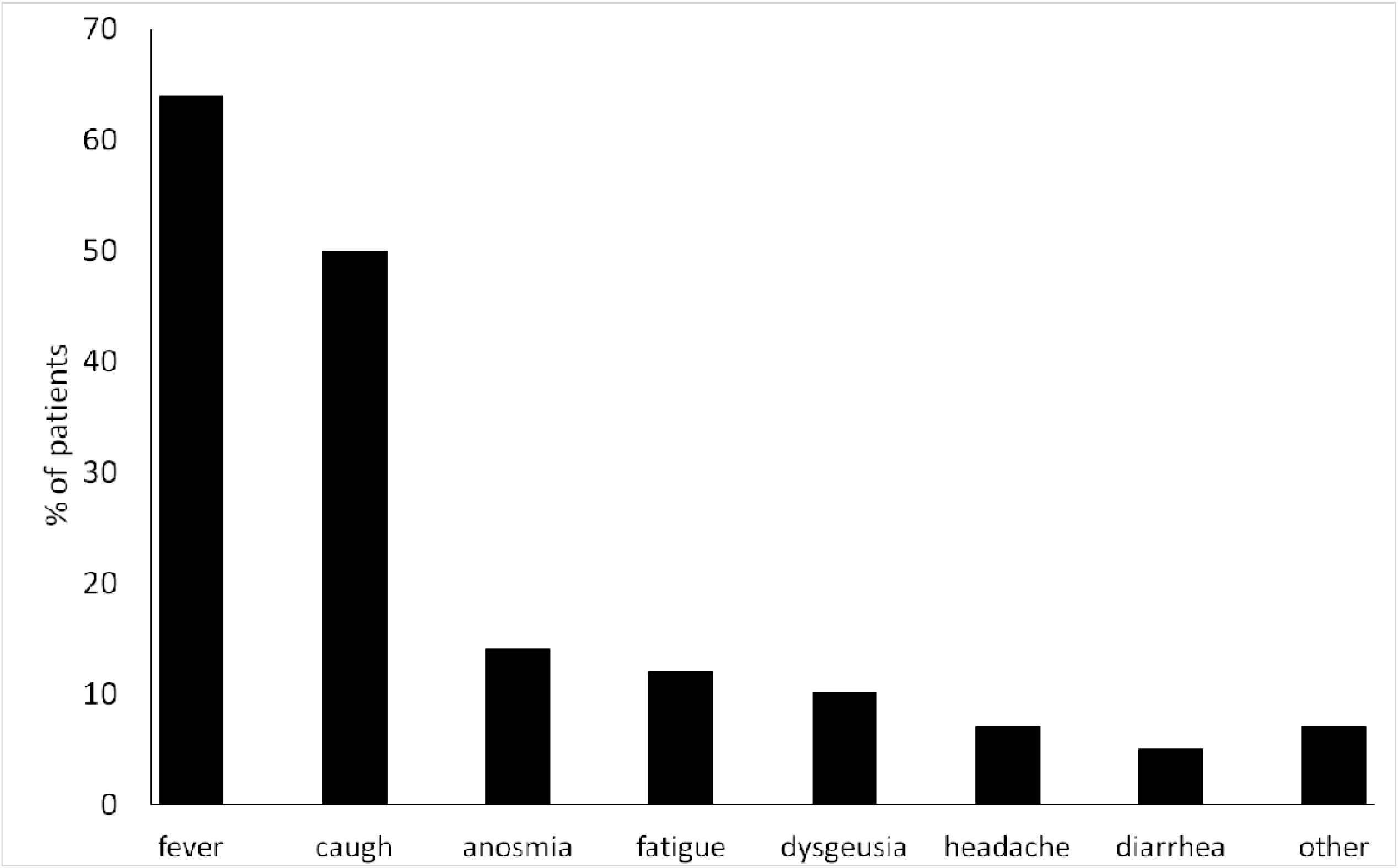
COVID-19 like symptoms reported by celiac patients during a lockdown period in the Milan area.

The ISMA (International Stress Management Association) Stress questionnaire demonstrated that 12% of the patients had a low probability of suffering from stress-related illness (score 0-4), 73% reported stress-related health, mental or physical problems, for which patients could benefit from counselling (score 5-13) and 15% showed the inclination to stress with unhealthy behaviours (score 14-25). No demographic difference was observed between CD patients who were or not suffering from stress related illness.

### Anti-SARS-CoV-2 antibodies and intestinal ACE2 receptor

109 (31%) CD patients (88 females, age at enrolment 62±13 years, age at CD diagnosis 33±13 years) underwent a serological detection of IgG and IgA against RBD and N proteins. Among them, the presence of at least one of the tested Ig was detected in 20 (18%) CD patients and in 42 (25%) control subjects (p=0.18). In the group of patients testing positive for anti-SARS-CoV-2 Ig, 16 (80%) developed anti-SARS-CoV-2 IgA while IgG antibodies were present in 15 (75%) individuals. Among them, 15 (75%) presented anti-RBD antibodies and 13 (65%) anti-N antibodies (figure 2A). In figure 2B, the OD of the anti-SARS-CoV-2 Ig of CD patients and controls are reported; notably, CD patients showed significantly reduced values of anti-N IgA compared to otherwise healthy individuals.

**Figure 2.**
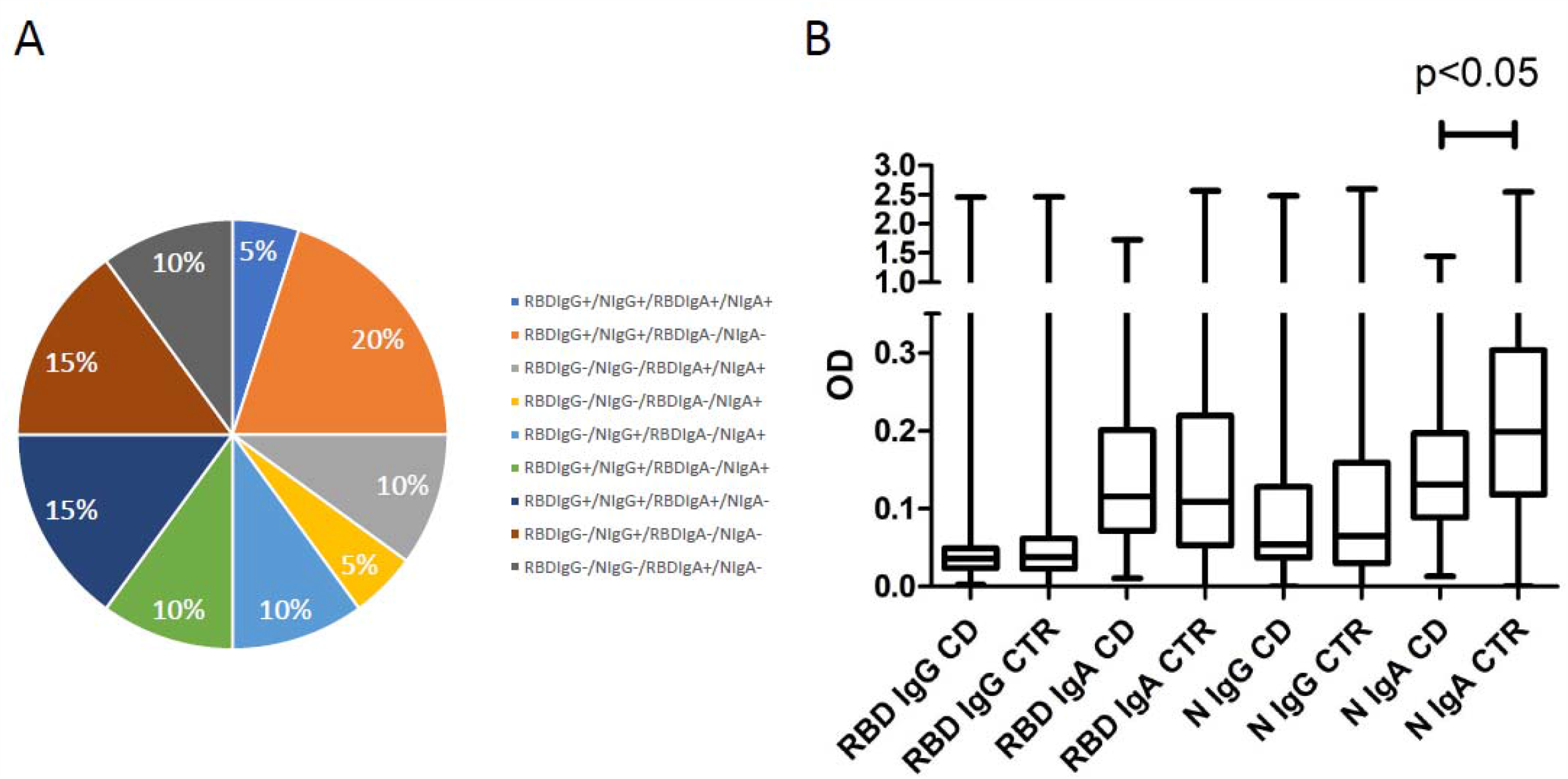
Distribution of the different Ig types against RBD and N proteins (A) and their values compared to the controls (B)

Among the 20 CD patients with at least one detectable Ig class in the serum, 9 (45%) were referred for COVID-19 like symptoms. Conversely, 17 (19%) individuals reported COVID-19 like symptoms albeit no anti-SARS-CoV-2 Ig were detected. In table 2, the clinical, serological and histological characteristics of the patients are reported. Twenty-seven (64%) of the 42 patients with COVID-19 symptoms underwent the serological test and 10 (37%) presented anti-SARS-CoV-2 Ig. CD patients with anti-SARS-CoV-2 Ig were asked to undergo a nasopharyngeal swab and one asymptomatic patient resulted in a positive result.

**Table 2.** Clinical and demographic characteristics of celiac patients with or without anti-SARS-CoV-2 Ig

|  | **Anti-SARS-CoV-2 antibodies** |  | **P** |
| --- | --- | --- | --- |
|  | **Positive** | **Negative** |  |
|  | **N=20** | **N=89** |  |
| **Sex, female (%)** | 16(80) | 72(81) | 1.0 |
| **Age at enrolment (years)** | 47±13 | 44±15 | 0.54 |
| **Age at diagnosis (years)** | 34±17 | 33±16 | 0.80 |
| **BMI (kg/cm<sup>2</sup>)** | 23.5±5.8 | 22.4±3.8 | 0.29 |
| **COVID-19 like symptoms n(%)** | 9(80) | 17(19) | 0.02 |
| **Positive anti-transglutaminase IgA (%)** | 5(25) | 17(19) | 0.54 |
| **Nonadherence to GFD and/or positive urinary GIP (%)** | 0(0) | 5(6) | 0.58 |
| **Duodenal atrophy (Marsh 3a, 3b, 3c) (%)** | 7(35) | 29(32) | 1.0 |
| **Refractory celiac disease (%)** | 0(0) | 2(2) | 1.0 |
| **Presence of an AI comorbidity (%)** | 2(10) | 17(19) | 0.51 |

At the tissue level, the ACE2 receptor was present on the luminal surface of the duodenal villi with no signs of atrophy with an immunohistochemical score of 133±73; conversely, when duodenal atrophy was present, ACE2 was weakly expressed on the enteric cells (figure 3).

**Figure 3.**
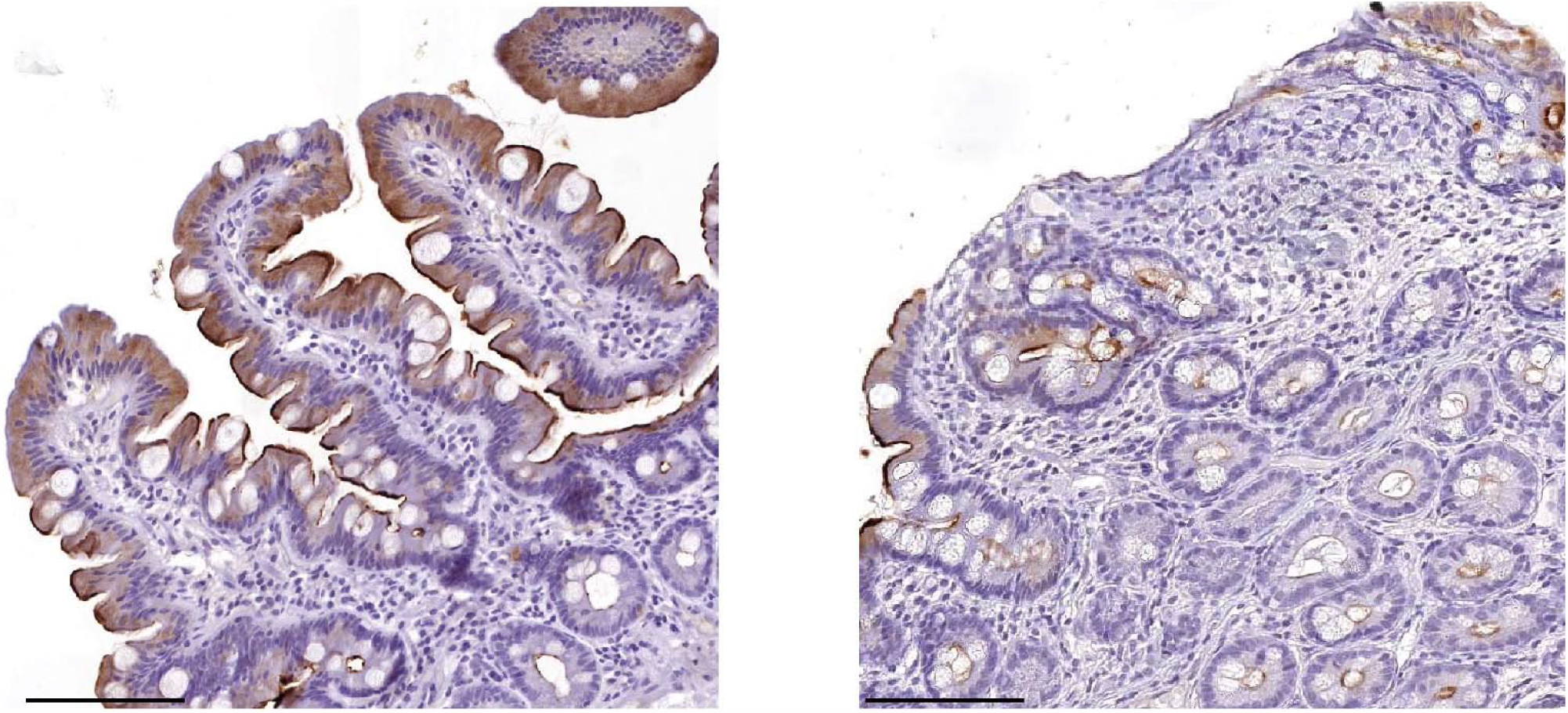
Representative images of ACE2 expression in a non-atrophic (inactive; A) and an active (B) CD patient. Scale bar, 100 µm.

## DISCUSSION

The present study represents the first attempt to investigate the effects of the SARS-CoV-2 pandemic on CD patients and their immunological response in case of viral infection. CD patients’ serological anti-SARS-CoV-2 Ig profile and percentage of positivity are similar to non-CD patients’ findings. Moreover, none of the most relevant CD biomarkers (tTGA, GFD adherence, duodenal atrophy) appears to influence the immune response in CD subjects, in spite of the expression of ACE2 receptor in the duodenum. In the investigated cohort of patients, symptoms related to SARS-CoV-2 infection correspond to those described in the literature with a single case of hospitalisation for mild respiratory distress. More than 80% of the CD patients present a stress related illness and the possible necessity of support due to the general pandemic scenario and lockdowns.

It should be underlined that the present findings are of particular relevance as they were obtained from an area heavily hit by the SARS-CoV-2 pandemic; in fact, all the patients attending the “Centre for Prevention and Diagnosis of Celiac Disease” of the Fondazione IRCCS Ca’ Granda Ospedale Maggiore Policlinico, live in the Milan-Bergamo Area (Northern Italy).[21] This factor is demonstrated by the high prevalence (around 20%) of patients presenting anti-SARS-CoV-2 Ig, independently by the presence or not of CD. Notably, 80% of patients with positive anti-SARS-CoV-2 serology reported the presence of symptoms, while 20% did not have any symptoms suggesting an asymptomatic contact. However, the number of symptomatic patients presented as negative at anti-SARS-CoV-2 Ig suggests that other factors could be involved in the patients’ clinical picture, or that the symptoms were not correlated to a SARS-CoV-2 infection. Unfortunately, during the first lockdown period, the execution of nasopharyngeal swabs was extremely limited, including at the hospitals, due to shortage of resources and, therefore, we are unable to present a PCR result to confirm or confute those findings.[21] However, serological testing has been used in recent months to monitor viral spreading in the general population and among individuals at increased risk of contracting the infection, including immunosuppressed patients.[22] Various commercial and in-house assays that utilise distinct viral antigens and detecting different antibody classes are currently available. Here, we used a test imported from the laboratory of Prof. Krammer[17,18] that received FDA approval for emergency use in the US. This test has an advantage over other types of tests to allow the simultaneous evaluation of anti-SARS-CoV-2 antibodies of different classes (IgA, IgG) raised against different types of viral antigens, including the receptor binding domain (RBD) of the Spike protein and the Nucleocapsid.[17–19] This type of assay was used to demonstrate that the magnitude of a neutralising Ab response may be associated with disease severity in COVID-19 patients and demonstrated its sensitivity also to evaluate the antibody response of paucisymptomatic individuals.[19] The specificity and sensitivity of the different assays greatly vary among kits taking into consideration the different techniques implemented (ELISA, CLIA, lateral flow) and the antigens used (Spike ectodomain, S1-S2 of the Spike, Spike RBD, Nucleocapsid). The assay used in this work showed an excellent performance in terms of specificity and sensitivity. Anti-RBD IgG showed a specificity and sensitivity of 97% and 95%, respectively, while the assay performed with the N protein values were 91% and 95%.[19] The performance of IgA detection was high for the RBD assay (91.5% specificity and 95% sensitivity), while it was slightly lower for the N protein (85% and 69%). These performances are in line with those published for both in-house and commercial assays.[22]

The IgA response against SARS-CoV-2 has been reported to be possibly associated with a mucosal immune response in the gut and lungs.[23] A recent paper suggested that IgA production might occur locally at the mucosal sites, possibly correlating with the viral load, the duration of the viral exposure and the virus entry route.[24] In line with this speculation, it has been shown recently that the highest levels of IgG and IgA antibodies against the Spike S1 domain were associated with severe disease.[25]

However, a multifactorial, CD genetic background is well known and strongly linked to HLA DQ2-DQ8 haplotypes. Several papers have focused on the identification of the possible predisposing genetic factors for either the development of the infection [26,27] or to a more severe clinical involvement.[28,29] In various cohorts, a significant HLA association was detected with the presence of SARS-CoV-2 infection, with class I and II molecules. However, among all studies, no significant association was detected with the DQ2 haplotype. As regards to the severity of the infection, although some studies identified some predisposing HLAs, the largest GWAS study did not report a significant role of the HLA region, whereas the AB0 blood group, and the 3p21.31 locus which harbours a cytokine/chemokine cluster were detected as significant.[28] Interestingly, a locus predisposing to celiac disease (CELIAC9) has been mapped to 3p21, but different genes are involved in SARS-CoV-2 and CD.[28] This is in agreement with the severity of a COVID-19 infection observed in the CD cohort reported herein, in which only one patient required hospitalisation.

CD pathologic cascade develops in the small bowel and duodenal atrophy represents the hallmark of the disease. We have found different levels of the ACE2 receptor in the mucosa of CD patients being very low in case of atrophy. Although we reported the first findings in CD, it was previously demonstrated that all the components of the renin-angiotensin system (RAS) are present in the small bowel mucosa. Furthermore, it was previously shown that ACE2 expression was significantly higher in the ileal tract than in the colonic tract of the intestine of patients without gastrointestinal disorders, and that its levels were decreased in the inflamed colonic biopsies from patients with inflammatory bowel disease (IBD). The role of the RAS system in the gut is to attenuate inflammatory states and to play anti-inflammatory, anti-fibrotic, and anti-proliferative actions in various tissues. In line with this, ACE2 knockout mice have increased susceptibility to colitis and an altered microbiota profile, compared to wild-type ones.[30] The fact that ACE2 is a constitutive protein of human small bowel is also supported by the fact that intestinal ACE2 is required for tryptophan absorption, an essential amino acid required for niacin production. Probably, a way by which SARS-CoV-2 elicits inflammation in the GI tract is related to the decrease of mucosal ACE2 presence after the virus entry into enteric cells.[31]

Because SARS-CoV-2 infects humans via the binding of the viral spike proteins with ACE2 protein on mucosal membranes, and the fact that SARS-CoV-2 spreading occurs also through the faeces,[32] we hypothesised that ACE2 levels in the small bowel mucosa of CD patients could predispose or protect these patients from severe COVID-19 disease. Gastrointestinal symptoms, including diarrhoea, occur in approximately 4%–20% of patients with COVID-19, and severe colitis has recently been described.

Deficiencies of both fat- and water-soluble vitamins have been documented in patients with CD.[33]

Although, the beneficial effects of a healthy diet are discussed in relation to the current COVID-19 pandemic and there are no known evidence-based therapeutics or treatment strategy included food advice available to prevent the incidence or severity of a COVID-19 infection, learning from previous research in relation to other viral infections, the nutritional status could play a significant role in patient outcomes.[34,35] Nowadays, there are various diets and nutrients that potentially exert anti-inflammatory and immunomodulatory effects on different diseases, including cardiovascular and lung diseases.[34] Notably, almost all CD patients referred had an optimal adherence to the GFD without problems in accessing to the GF products.

Notably, none of the routinely evaluated CD biomarkers (tTGA, duodenal atrophy, GFD adherence) influenced symptoms and/or Ig response. Based on our results and the study limits, we have found a CD serological anti-SARS-CoV-2 response similar to what is present in healthy controls. Moreover, CD patients reported an optimal adherence to the GFD and anti-viral lifestyle, although a relevant grade of stress is present, suggesting centres to offer a telehealth service.[36] The symptomatic profile and the absence of severe events, albeit in a high incidence scenario, do not support the presence of a different prognosis of SARS-CoV-2 infection in CD.

## Supporting information

strobe

## Data Availability

Data are not available

## Contributors

LE, LR and MV participated in the study conception, patient recruitment, data collection, statistical analysis, writing of the manuscript and the final approval of the manuscript. AC, AS and VL participated in the patient recruitment, phone calls and data collection. FF executed the laboratory tests and provided the control group. VV performed and analysed the immunohistochemistry. BDO evaluated the stress questionnaires. LS, DSS and LD thoroughly revised and finally approved the manuscript.

## Patient and public involvement

The patients were involved in the design and dissemination plans of this research. Refer to the Methods section for further details.

## Patient consent for publication

Not required.

## Ethics approval

Ethical approval for the study was obtained from the “Comitato Etico Milano Area 2” (reference number 458_2020).

## Provenance and peer review

Not commissioned; externally peer reviewed.

## Data availability statement

No data available.

## REFERENCES

1. Sun K, Chen J, Viboud C. Early epidemiological analysis of the coronavirus disease 2019 outbreak based on crowdsourced data: a population-level observational study. Lancet Digit Heal 2020;7500. doi:10.1016/S2589-7500(20)30026-1

2. Huang C, Wang Y, Li X, et al. Clinical features of patients infected with 2019 novel coronavirus in Wuhan, China. Lancet 2020;395:497–506. doi:10.1016/S0140-6736(20)30183-5

3. Guan W-J, Ni Z-Y, Hu Y, et al. Clinical Characteristics of Coronavirus Disease 2019 in China. N Engl J Med 2020;:1–13. doi:10.1056/NEJMoa2002032

4. Chen N, Zhou M, Dong X, et al. Epidemiological and clinical characteristics of 99 cases of 2019 novel coronavirus pneumonia in Wuhan, China: a descriptive study. Lancet 2020;395:507–13. doi:10.1016/S0140-6736(20)30211-7

5. Faye AS, Lee KE, Laszkowska M, et al. Risk of Adverse Outcomes in Hospitalized Patients with Autoimmune Disease and COVID-19: A Matched Cohort Study from New York City. J Rheumatol 2020;:jrheum.200989. doi:10.3899/jrheum.200989

6. Elli L, Scaramella L, Lombardo V, et al. Refractory celiac disease and COVID-19 outbreak: Findings from a high incidence scenario in Northern Italy. Clin. Res. Hepatol. Gastroenterol. 2020;44:e115–20. doi:10.1016/j.clinre.2020.07.026

7. Zhen J, Stefanolo JP, Temprano MP, et al. THE RISK OF CONTRACTING COVID-19 IS NOT INCREASED IN PATIENTS WITH CELIAC DISEASE. Clin Gastroenterol Hepatol Published Online First: October 2020. doi:10.1016/j.cgh.2020.10.009

8. Gu J, Han B, Wang J. COVID-19: Gastrointestinal manifestations and potential fecal-oral transmission. Gastroenterology. 2020;E-pub ahea. doi:10.1053/j.gastro.2020.02.054

9. Lu J, Sun PD. High affinity binding of SARS-CoV-2 spike protein enhances ACE2 carboxypeptidase activity. J Biol Chem 2020;:jbc.RA120.015303. doi:10.1074/jbc.RA120.015303

10. Garg M, Royce SG, Lubel JS. Letter: intestinal inflammation, COVID-19 and gastrointestinal ACE2 - exploring RAS inhibitors. Aliment Pharmacol Ther Published Online First: 6 May 2020. doi:10.1111/apt.15814

11. Elli L, Ferretti F, Orlando S, et al. Management of celiac disease in daily clinical practice. Eur. J. Intern. Med. 2019;61:15–24. doi:10.1016/j.ejim.2018.11.012

12. Elli L, Villalta D, Roncoroni L, et al. Nomenclature and diagnosis of gluten-related disorders: A position statement by the Italian Association of Hospital Gastroenterologists and Endoscopists (AIGO). Dig Liver Dis 2017;49. doi:10.1016/j.dld.2016.10.016

13. Elli L, Barisani D, Vaira V, et al. How to manage celiac disease and gluten-free diet during the COVID-19 era: proposals from a tertiary referral center in a high-incidence scenario. BMC Gastroenterol 2020;20:387. doi:10.1186/s12876-020-01524-4

14. Elli L, Rimondi A, Scaramella L, et al. Endoscopy during the Covid-19 outbreak: experience and recommendations from a single center in a high-incidence scenario. Dig Liver Dis 2020;52:606–12. doi:10.1016/j.dld.2020.04.018

15. Oberhuber G, Granditsch G, Vogelsang H. The histopathology of coeliac disease. Eur J Gastroenterol Hepatol 1999;11:1185. doi:10.1097/00042737-199910000-00019

16. Gligor LJerban, MozoLJ I. Indicators of smartphone addiction and stress score in university students. Wien Klin Wochenschr 2019;131:120–5. doi:10.1007/s00508-018-1373-5

17. Amanat F, Stadlbauer D, Strohmeier S, et al. A serological assay to detect SARS-CoV-2 seroconversion in humans. Nat Med 2020;26:1033–6. doi:10.1038/s41591-020-0913-5

18. Stadlbauer D, Amanat F, Chromikova V, et al. SARS-CoV-2 Seroconversion in Humans: A Detailed Protocol for a Serological Assay, Antigen Production, and Test Setup. Curr Protoc Microbiol 2020;57. doi:10.1002/cpmc.100

19. Bruni M, Cecatiello V, Diaz-Basabe A, et al. Persistence of Anti-SARS-CoV-2 Antibodies in Non-Hospitalized COVID-19 Convalescent Health Care Workers. J Clin Med 2020;9:3188. doi:10.3390/jcm9103188

20. Vaira V, Gaudioso G, Laginestra MA, et al. Deregulation of miRNAs-cMYC circuits is a key event in refractory celiac disease type-2 lymphomagenesis. Clin Sci 2020;134:1151–66. doi:10.1042/CS20200032

21. Remuzzi A, Remuzzi G. COVID-19 and Italy: what next? Lancet. 2020;395:1225–8. doi:10.1016/S0140-6736(20)30627-9

22. Caini S, Bellerba F, Corso F, et al. Meta-analysis of diagnostic performance of serological tests for SARS-CoV-2 antibodies up to 25 April 2020 and public health implications. Eurosurveillance 2020;25. doi:10.2807/1560-7917.ES.2020.25.23.2000980

23. Seow J, Graham C, Merrick B, et al. Longitudinal observation and decline of neutralizing antibody responses in the three months following SARS-CoV-2 infection in humans. Nat Microbiol 2020;5. doi:10.1038/s41564-020-00813-8

24. Padoan A, Sciacovelli L, Basso D, et al. IgA-Ab response to spike glycoprotein of SARS-CoV-2 in patients with COVID-19: A longitudinal study. Clin Chim Acta 2020;507:164–6. doi:10.1016/j.cca.2020.04.026

25. Carsetti R, Zaffina S, Piano Mortari E, et al. Spectrum of innate and adaptive immune response to SARS CoV 2 infection across asymptomatic, mild and severe cases; a longitudinal cohort study. Concetta Quintarelli 2020;13:21. doi:10.1101/2020.06.22.20137141

26. Wang W, Zhang W, Zhang J, et al. Distribution of <scp>HLA</scp> allele frequencies in 82 Chinese individuals with coronavirus disease-2019 (COVID-19). HLA 2020;96:194–6. doi:10.1111/tan.13941

27. Novelli A, Andreani M, Biancolella M, et al. HLA allele frequencies and susceptibility to COVID-19 in a group of 99 Italian patients. HLA Published Online First: 2020. doi:10.1111/tan.14047

28. de Sousa E, Ligeiro D, Lérias JR, et al. Mortality in COVID-19 disease patients: Correlating the association of major histocompatibility complex (MHC) with severe acute respiratory syndrome 2 (SARS-CoV-2) variants. Int J Infect Dis 2020;98:454–9. doi:10.1016/j.ijid.2020.07.016

29. Lorente L, Martín MM, Franco A, et al. HLA genetic polymorphisms and prognosis of patients with COVID-19. Med Intensiva Published Online First: 2020. doi:10.1016/j.medin.2020.08.004

30. Garg M, Christensen B, Lubel JS. Gastrointestinal ACE2, COVID-19 and IBD: Opportunity in the Face of Tragedy? Gastroenterology. 2020;159:1623-1624.e3. doi:10.1053/j.gastro.2020.04.051

31. Xiao F, Tang M, Zheng X, et al. Evidence for gastrointestinal infection of SARS-CoV-2. Gastroenterology 2020;E-pub ahea.

32. Du M, Cai G, Chen F, et al. Multiomics Evaluation of Gastrointestinal and Other Clinical Characteristics of COVID-19. Gastroenterology 2020;158:2298-2301.e7. doi:10.1053/j.gastro.2020.03.045

33. Theethira TG, Dennis M. Celiac disease and the gluten-free diet: Consequences and recommendations for improvement. Dig Dis 2015;33:175–82. doi:10.1159/000369504

34. Zabetakis I, Lordan R, Norton C, et al. Covid-19: The inflammation link and the role of nutrition in potential mitigation. Nutrients. 2020;12. doi:10.3390/nu12051466

35. Mercola J, Grant WB, Wagner CL. Evidence Regarding Vitamin D and Risk of COVID-19 and Its Severity. Nutrients 2020;12:3361. doi:10.3390/nu12113361

36. Siniscalchi M, Zingone F, Savarino EV, et al. COVID-19 pandemic perception in adults with celiac disease: an impulse to implement the use of telemedicine. Dig Liver Dis 2020;0. doi:10.1016/j.dld.2020.05.014

